# Clinical Trial Components of NIH Career Development Awards: Supporting the Nation’s Future Trialists

**DOI:** 10.64898/2026.09.18.26363419

**Authors:** Catherine G. Derington, Lilia Cervantes, Larry A. Allen, Katy E. Trinkley, Amy G. Huebschmann, P. Michael Ho

**Affiliations:** Department of Medicine, School of Medicine, University of Colorado Anschutz, Aurora, CO, USA; Adult & Child Center for Outcomes Research & Delivery Science, University of Colorado Anschutz, Aurora, CO, USA; Department of Family Medicine, School of Medicine, University of Colorado Anschutz, Aurora, CO, USA; Institute for Health Research, Kaiser Permanente Colorado, Aurora, CO, USA

## Abstract

In 2027, the National Institutes of Health (NIH) will no longer support costs associated with clinical trials (CT) for career development (K) awards. NIH’s CT definition encompasses many types of research, from early-stage pilot work through definitive studies. The types and sizes of trials K awardees conduct have not been described, which could inform ongoing policy changes relevant to the next generation of clinical trialists. We conducted a cross-sectional analysis of NIH RePORTER data to characterize the types and sizes of CTs supported by NIH K01, K08, and K23 awards funded in fiscal years 2024-2025 under a CT Required or CT Optional funding opportunity announcement (FOA), with non-missing abstract text. A rule-based classification scheme applied to free-text abstracts categorized each award into 5 mutually exclusive trial types: pilot/feasibility/proof-of-concept, efficacy/effectiveness, trial confirmed with type unclear, other (mechanistic, ancillary, or phase I/II), or no trial-specific language identified. Trial size was assessed among abstracts reporting an explicit, manually verified sample size. Of 1336 unique awards funded under CT Required (n=1051) or CT Optional (n=285) FOAs, 815 (61.0%) were classified as pilot, feasibility, or proof-of-concept studies, and 55 (4.1%) as efficacy or effectiveness studies; 204 (15.3%) had a confirmed trial of unclear type, 41 (3.1%) were other, and 221 (16.5%) had no trial-specific language. Among CT Required awards, 730 (69.5%) were pilot, feasibility, or proof-of-concept work. Among 273 awards (20.4%) with a verified sample size, the median was 60 participants (IQR, 44-80), with 236 (86.4%) enrolling ≤100 participants. Given these findings that CT within NIH K awards are generally small and early-stage, NIH’s decision to no longer support trial costs may adversely affect the scientist development pipeline.

## Introduction

In clinical research, clinical trials (CT) are conducted to test whether an intervention is safe and effective in humans. The United States’ National Institutes of Health (NIH) defines a CT as any research study that prospectively assigns human subjects to interventions to evaluate health-related biomedical or behavioral outcomes.^1^ This broad definition encompasses a wide spectrum of trial types and designs, from formative pilot trials through definitive trials that demonstrate efficacy or effectiveness.^2^ Each type and size of trial funded by NIH serves a specific, methodologically necessary purpose in the translational research pipeline. As CTs can be complex and resource-intensive, early-career scientists can receive CT training through mentored career development (K) awards, which allows them to compete for larger grants later to conduct definitive trials.

On August 12, 2026, NIH announced that it will eliminate “Independent Clinical Trial Required” funding opportunities for K awards; awardees will instead need to secure CT funding elsewhere (NOT-OD-26-098), with unclear plans for how CT training will be provided to support the career development of the next generation of clinical trialists.^3^ A companion Request for Information seeks comment on 2 replacement funding models,^4^ but there are no empirical data describing the types or sizes of CTs currently supported by K awards. These data are critical to understand downstream impacts of eliminating CTs from K mechanisms. For example, if most K awards support early-stage, pilot work, then eliminating this funding would defund the same type of early-stage work that NIH supports elsewhere through mechanisms like R21 and R34 grants, and would sever the continuity between the investigator who conducts pilot work and the definitive trial that follows.

To fill this gap and inform this ongoing policy discussion, we characterized the types and sizes of CTs funded in NIH K awards.

## Methods

On September 1, 2026, we queried NIH RePORTER for K01, K08, and K23 awards funded in fiscal years 2024 and 2025. We de-duplicated awards by project number, and restricted the sample to awards funded under CT Required or CT Optional funding opportunity announcements (FOA).

To characterize the CT type of each award, we developed a rule-based text classification scheme and applied it to the free-text project abstract (**eMethods**). Classification used keyword and phrase patterns (regular expressions) applied in priority order and refined iteratively. The final scheme had 5 mutually exclusive categories: (1) pilot, feasibility or proof-of-concept; (2) efficacy or effectiveness (including hybrid implementation designs); (3) trial confirmed, type unclear; (4) other (mechanistic, ancillary, or regulatory phase-labeled trials); and (5) no trial-specific language identified.

We validated the automated classification against manual review of 300 randomly sampled abstracts over 3 rounds of pattern refinement; agreement was substantial (Cohen κ=0.76; 86.7% raw agreement; **eMethods**). For awards with a manually verified sample size, we characterized the size of the CT reported in the abstract, excluding enrollment counted in group-level units (eg, “clinics”) unless the unit specified a fixed number of individuals (eg, dyads, triads). The classification code, the final pattern set (**eTable 1**), and adjudicated overrides (**eTable 2**) are available in the Online Supplement.

Descriptive statistics (i.e., median (IQR) for continuous variables and number (percent) for categorical variables) were used for analyses overall and by FOA type (CT Required vs. Optional). The abstract text classification scheme (pattern development, iterative refinement, and application) and descriptive statistical analyses were performed with assistance from Claude version Sonnet 5 (Anthropic), an AI large language model, used on September 1-8, 2026. The study team reviewed all AI-generated code, classifications, and analytic output, manually overrode classifications where indicated, and takes full responsibility for the accuracy and integrity of the reported methodology and results.

## Results

The NIH RePORTER query returned 7,238 records representing 4,162 unique awards with available abstracts; of these, n=1,336 (32.1%) were funded under CT Required (N=1,051) or CT Optional (n=285) FOAs. The most common activity code was K23 (776 [58.0%]), followed by K01 (375 [28.1%]), then K08 (185 [13.8%]) (**Table 1**).

**Table 1.**
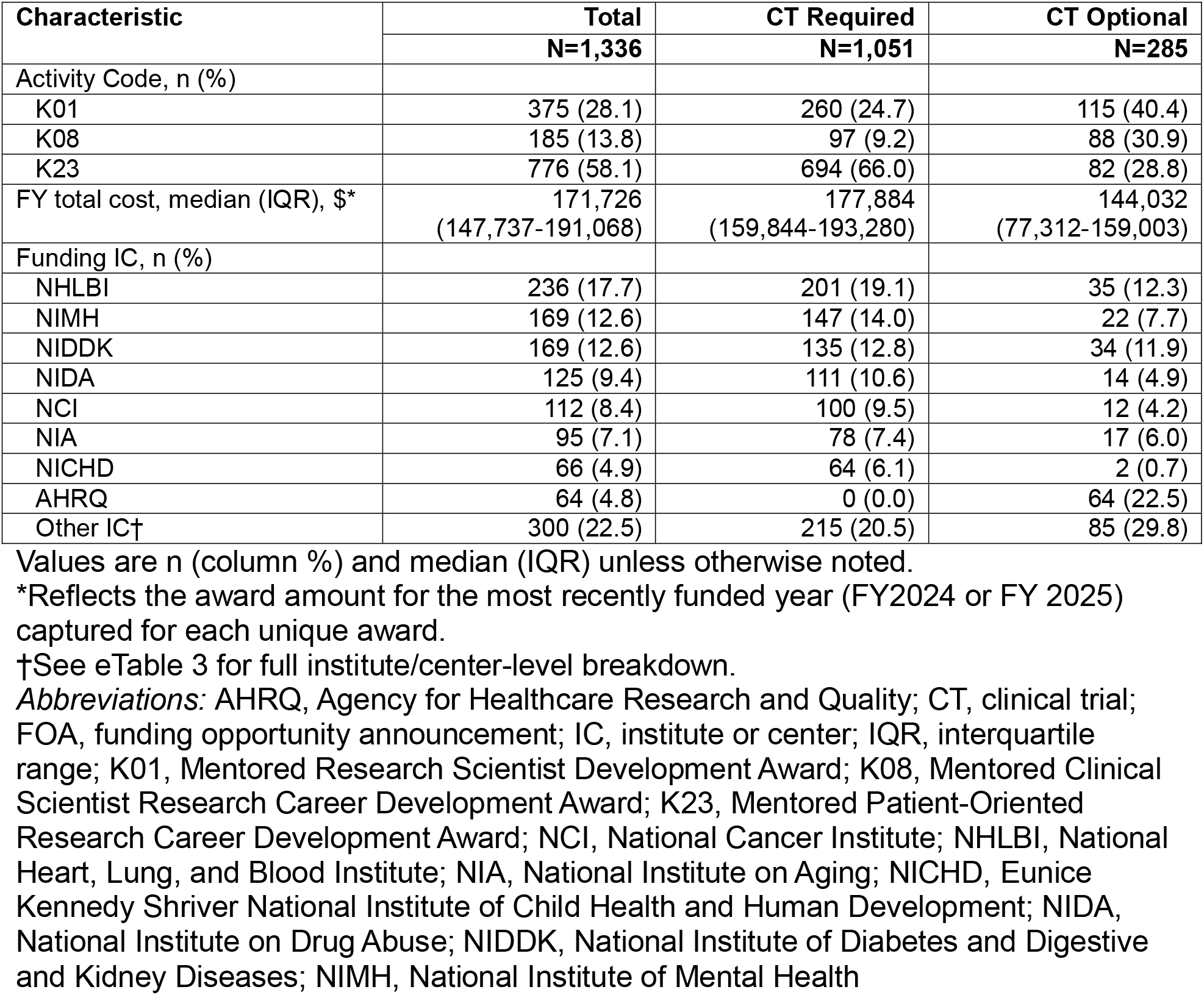
Characteristics of K01/K08/K23 Awards Funded Under Clinical Trial Required and Clinical Trial Optional FOAs, FY2024–2025.

Overall, n=815 of 1,338 (61.0%) were classified as pilot, feasibility, or proof-of-concept work; 55 (4.1%) described a definitive efficacy or effectiveness trial (**Table 2**). Among CT Required awards, n=730 of 1,051 (69.5%) were pilot, feasibility, or proof-of-concept and 48 (4.6%) described a definitive efficacy or effectiveness trial.

**Table 2.**
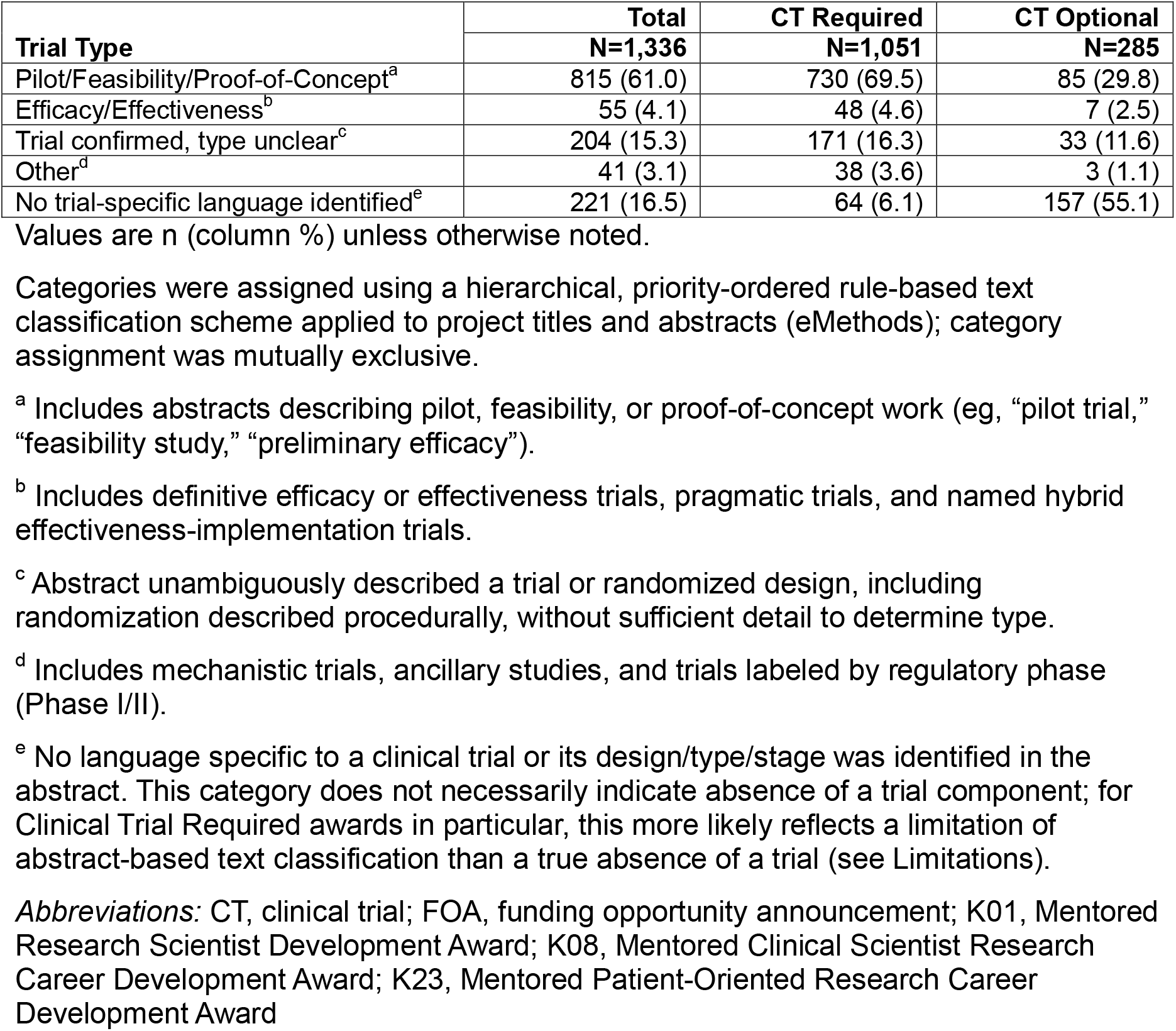
Characteristics of K01/K08/K23 Awards Funded Under Clinical Trial Required and Clinical Trial Optional FOAs, FY2024–2025.

A trial sample size could be extracted and manually verified from the abstract for n=273 of 1,336 awards (20.4%). Among these, the median proposed sample size was 60 (IQR, 44-80; range, 12-900); n=236 (86.4%) planned to enroll ≤100 participants (**Figure 1**).

**Figure 1.**
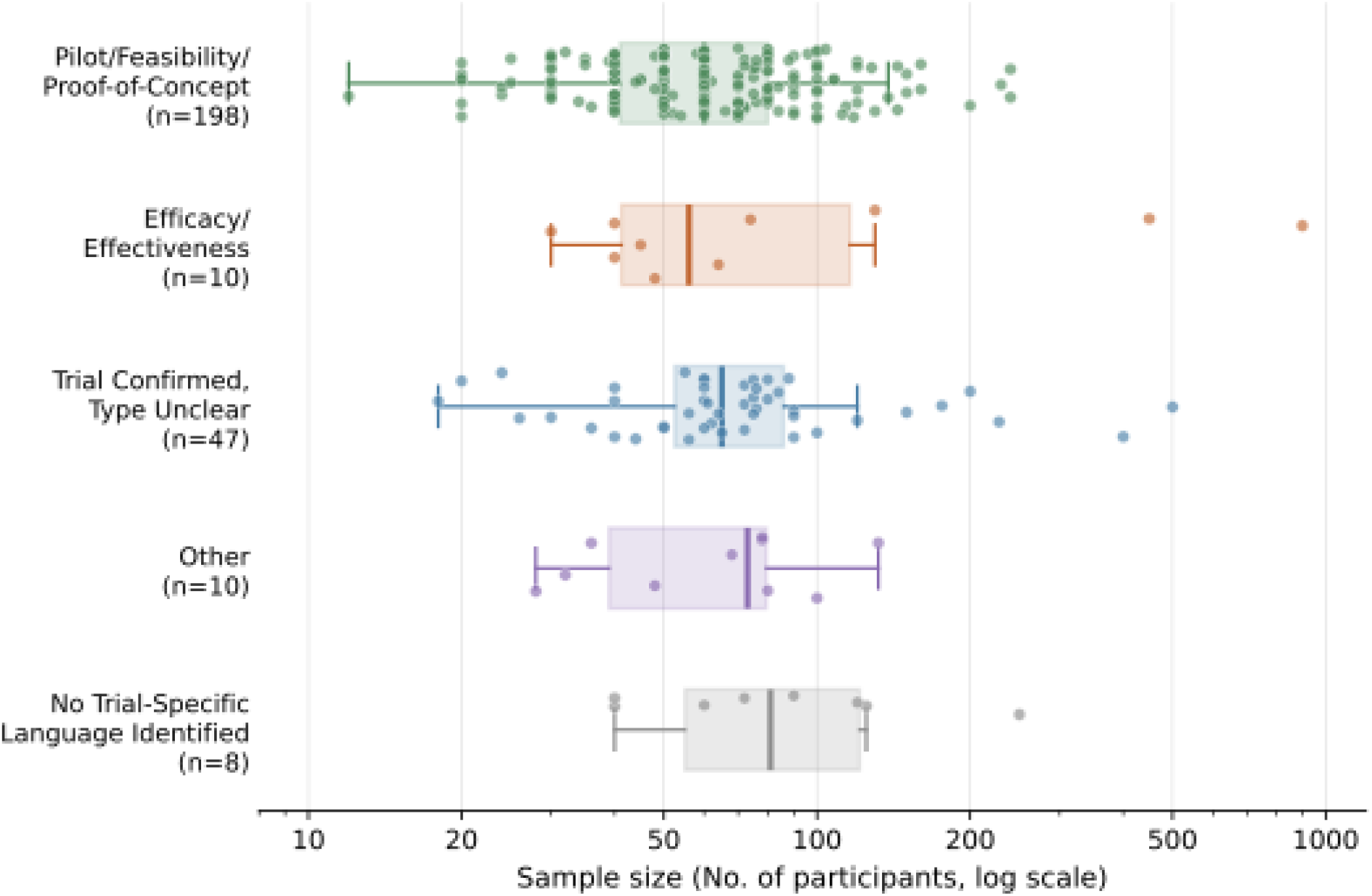
Clinical Trial Sample Size Among K01, K08, and K23 Awards funded by NIH in FY 2024-2025, by Trial Type. Each point represents 1 award (N=273); boxes show median and interquartile range (IQR); whiskers extend to the most extreme value within 1.5×IQR of the box, with values beyond this range plotted as individual points rather than excluded. Sample sizes were manually verified from project abstracts among the subset of 1,336 awards funded under Clinical Trial Required or Clinical Trial Optional funding opportunity announcements (FY2024-2025) for which an interpretable sample size was reported in the abstract text (n=273 [20.4%]). Categories were assigned using a hierarchical, rule-based text classification scheme (eMethods; eTable). Median (IQR; range), by category: Pilot/Feasibility/Proof-of-Concept, 60 (41-80; 12-240); Efficacy/Effectiveness, 56 (41-116; 30-900); Trial Confirmed, Type Unclear, 65 (52-86; 18-500); Other, 73 (39-80; 28-132); No Trial-Specific Language Identified, 81 (55-121; 40-250).

## Discussion

These findings suggest that CTs proposed by K01, K08, and K23 awards are predominantly small-scale, early-stage pilot work intended to build the infrastructure and training needed for a future definitive trial. Pilot and feasibility work occupies a well-established, methodologically necessary stage in the translational research pipeline: testing whether an intervention is feasible, acceptable, and implementable before committing the substantially larger resources required for a definitive CT.^5^ Without a mechanism to conduct pilot work as part of a K award, investigators risk spending protected time and public funds on interventions that later prove infeasible, unacceptable, or unimplementable at scale. In parallel, the nation risks losing a competent workforce of clinical trialists.

There is no “ideal” sample size for a pilot CT, which are intended to be small in nature while mirroring the future definitive CT design so that the investigator can build infrastructure and relationships and solidify essential components of the trial, like recruitment, retention/attrition, consent, treatment fidelity and adherence, or outcome measurement procedures.^2,6^ The small scale of K award trials in the current study (median n=60) reflects these objectives. One consideration may be modifying NIH’s K models and regulatory framework to differentiate pilot CTs from their traditional CT definition, unchanged since 2014, which does not currently distinguish CTs by type, scale, or risk.^1^

The funding models proposed in the companion Request for Information preserve K awardees’ ability to lead a trial but not NIH’s direct financial support for doing so, shifting risk onto early-stage investigators and mentors. Because supplemental funding is inherently uncertain, this approach leaves practical questions unresolved if funding is unsuccessful or lapses. The phased K99/R00-based transition model also warrants scrutiny for clinical investigators, since this mechanism has primarily supported PhD applicants in basic science and may require modified review criteria or panels.^7^ These uncertainties fall on junior investigators already navigating declining success rates (from 34%-40% in FY2014 to 23%-31% in FY2025).^8^

This study’s limitations include risk for errors from the rule-based classification scheme; reliance on abstract text alone, which may omit details; and a small subset with extractable sample size that may not generalize. Additionally, no clinical trial–specific language was identified for 6.1% of CT Required and 55.1% of CT Optional awards, likely reflecting undercapture for Clinical Trial Required awards and a true absence of a trial component for Clinical Trial Optional awards. Restriction to FY2024-2025 does not capture trends over time.

As NIH finalizes its approach to training the next generation of clinical trialists, replacement funding mechanisms should be calibrated to the work K awardees actually do and need to conduct. Removing or complicating K funding for pilot and feasibility trials risks discontinuity across the research enterprise.

## Data Sharing Statement

Will data be shared (including data dictionaries)? Yes.

What data will be shared? The deduplicated, classified analytic dataset (award-level data derived from NIH RePORTER, including funding opportunity announcement clinical trial designation and clinical trial stage/design classification) and the full analytic code, including the text classification pattern set (regular expressions) and scripts used to generate Tables 1 and 2.

What other documents will be available? The classification scheme documentation (eTable, mapping each pattern-matching category to its corresponding Table 2 category) and the full institute/center funding breakdown (eTable).

When will data be available (start and end dates)? Beginning immediately upon publication, with no end date.

By what access criteria will data be shared (including with whom, for what types of analyses, and by what mechanism)? NIH RePORTER data are publicly available to anyone, for any purpose, without requiring a proposal or investigator agreement, via https://reporter.nih.gov/.

## Data Availability

All data produced are available online at reporter.nih.gov

https://www.reporter.nih.gov

## Acknowledgements

The authors used Claude Sonnet 5 (Anthropic; September 1-8, 2026) to assist with manuscript preparation, including drafting and revising text and verifying arithmetic and percentage calculations reported in the text. The authors reviewed, verified, and take full responsibility for the accuracy and integrity of all content in the manuscript.

