## Supplemental Methods and Tables for "Clinical Trial Components of NIH Career Development Awards: Supporting the Nation’s Future Trialists"

**eMethods - Development of the rule-based text classification scheme**

An initial pattern set was constructed from terms *a priori* expected to signal trial type or design (eg, "pilot," "feasibility," "randomized clinical trial"). This initial classification was iteratively refined through manual review of a random sample of abstracts assigned to each category, with particular attention to the residual category of abstracts containing no detectable trial-specific language. Refinement addressed three recurring sources of misclassification: (1) morphological and syntactic variants not captured by the initial patterns (eg, verb forms such as "piloted" or "pilot-tested," and phrases separated by intervening descriptive text, such as "pilot a single-center study"); (2) systematic missing-space artifacts in the exported abstract text (eg, "thefeasibility") that broke word-boundary matching and required tolerance for character-level concatenation; and (3) qualifying language altering the interpretation of otherwise definitive-sounding terms (eg, "preliminary efficacy," which was reclassified from efficacy/effectiveness to pilot/feasibility/proof-of-concept, consistent with its common usage in career development award abstracts to describe underpowered, exploratory analyses).

Categories were assigned using a mutually exclusive, priority-ordered hierarchy, such that more specific or definitive design/stage language (eg, a named hybrid effectiveness-implementation design) took precedence over generic or procedurally-described language (eg, an isolated aim to "assess feasibility") when both were present in the same abstract, to avoid a less specific descriptor superseding a more informative one elsewhere in the same text. Classification was performed using a hierarchical, priority-ordered set of keyword and phrase patterns (regular expressions) developed iteratively rather than a predefined fixed lexicon, reflecting the exploratory nature of characterizing how investigators describe trial design and stage in unstructured text.

The first author manually reviewed 3 sequential random samples of 100 abstracts each (N=300 total) against the automated classification, documenting discrepancies with a rationale after each round; each round informed further refinement of the classification pattern set prior to the next round of review. Agreement between the automated classification and manual review was assessed using Cohen κ, calculated for each round and cumulatively. In 34 cases, the automated classification was overridden where the reviewer's holistic reading of the full abstract diverged from the automated keyword match. Agreement between automated classification and manual review improved across successive rounds of pattern refinement (round 1: κ=0.69; round 2: κ=0.74; round 3: κ=0.83), with substantial overall agreement across all 300 reviewed abstracts (κ=0.76; 86.7% raw agreement).

The final scheme comprised 5 mutually exclusive categories: (1) *Pilot/Feasibility/Proof-of-Concept*, denoting early-stage testing; (2) *Efficacy/Effectiveness*, denoting definitive testing of an intervention's clinical benefit, including hybrid effectiveness-implementation designs; (3) *Trial Confirmed, Type Unclear*, denoting abstracts that unambiguously described a trial or “randomized” design without sufficient additional detail to determine type; (4) *Other*, comprising mechanistic, ancillary, and regulatory phase-labeled (Phase I/II) trials, which describe trial purpose or regulatory status rather than developmental stage; and (5) *No Trial-Specific Language Identified*, for abstracts in which no language specific to a CT or its design/type/stage was identified.

Among awards with a manually verified sample size, we described the sample size reported in the abstract text; sample sizes were extracted and independently verified against the original abstract, using the maximum enrollment figure when an abstract reported sizes for multiple trial aims. Sample sizes referring to a group-level unit of enrollment (eg, clinics, families, health systems) were not counted unless the unit was a specific, defined multi-person group (eg, dyads, triads).

### eTable 1. Final Pattern Set Classification Scheme for Clinical Trial Stage/Design Language in K01/K08/K23 Abstracts.

| Category (V3) | Definition | Representative trigger language | Corresponding Table 2 Category |
|---|---|---|---|
| Pilot | Early-stage or exploratory testing of an intervention, explicitly labeled as a pilot, or described using pilot-related verb forms. Excludes prior/existing pilot-trial data being analyzed. Broadened in V3 to include a general "pilot the ___" construction regardless of target noun. [V3] | *"pilot trial", "pilot study", "piloted", "pilot test(ed/ing)", "pilot the [tools/implementation/etc.]"* | Pilot/Feasibility/Proof-of-Concept |
| Feasibility | Early-stage testing framed around feasibility, acceptability, or preliminary efficacy/effectiveness. MOVED UP in V3 priority order (now checked before Hybrid/Pragmatic/Phase/Effectiveness/Efficacy) so a "feasibility trial" qualifier is not missed when it co-occurs with hybrid/effectiveness language. Now also excludes prior/existing feasibility-study mentions. [V3] | *"feasibility trial/study", "feasibility and effectiveness", "hybrid…feasibility trial", "preliminary efficacy/effectiveness"* | Pilot/Feasibility/Proof-of-Concept |
| Proof-of-Concept | Early-stage demonstration that an approach is viable in principle. | *"proof-of-concept"* | Pilot/Feasibility/Proof-of-Concept |
| Efficacy | Definitive testing of clinical benefit, not qualified by "preliminary." Excludes future-trial mentions. | *"efficacy trial", "efficacy study"* | Efficacy/Effectiveness |
| Effectiveness | Definitive testing of real-world clinical benefit, not qualified by "preliminary." Excludes future-trial mentions. | *"effectiveness trial", "effectiveness study"* | Efficacy/Effectiveness |
| Efficacy (broad aims-phrasing) | Low-priority catch-all: an aim to test/assess/determine the efficacy of an intervention. In V3, benefits from the general "of" missing-space fix, which previously caused some genuine matches (e.g., "efficacy of[ ]oral") to be missed entirely. [V3] | *"[assess/determine/test] the efficacy of…"* | Efficacy/Effectiveness |
| Effectiveness (broad aims-phrasing) | Low-priority catch-all: an aim to test/assess/determine the effectiveness of an intervention. | *"[assess/determine/test] the effectiveness of…"* | Efficacy/Effectiveness |
| Hybrid Effectiveness-Implementation | A named hybrid effectiveness-implementation design. Excludes future-trial mentions. | *"hybrid type 1/2/3 effectiveness-implementation trial"* | Efficacy/Effectiveness |
| Pragmatic | A trial explicitly described as pragmatic. Excludes future-trial mentions. | *"pragmatic trial"* | Efficacy/Effectiveness |
| Randomized/Controlled Trial (stage unspecified) | Randomized/controlled design confirmed, stage unspecified. Excludes future-trial mentions. | *"randomized controlled trial", "RCT"* | Trial Confirmed, Stage Unclear |
| Randomized design described (no 'trial'/'RCT' label used) | Randomization described procedurally, without the words "trial" or "RCT." | *"will be randomly assigned to…"* | Trial Confirmed, Stage Unclear |
| Trial mentioned (no stage/design descriptor) | The word "trial" appears, with no further design/stage detail. | *"trial" (generic, catch-all)* | Trial Confirmed, Stage Unclear |
| Generic 'clinical trial' (no specific stage descriptor) | "Clinical trial" appears generically, no further detail. | *"clinical trial" (generic, catch-all)* | Trial Confirmed, Stage Unclear |
| Mechanistic | Trial/study purpose is elucidating a mechanism. Broadened in V3 beyond the literal "mechanistic trial" phrase to include "mechanistic role/understanding/insight/characterization/link(s)/pathway." Now also excludes future-trial mentions (previously unmasked in V2). [V3] | *"mechanistic trial", "mechanistic role of X", "gain mechanistic understanding"* | Other |

| Category (V3) | Definition | Representative trigger language | Corresponding Table 2 Category |
|---|---|---|---|
| Ancillary | A sub-study within an existing parent trial. | *"ancillary trial/study"* | Other |
| Phase I/II (regulatory) | Trial labeled with a regulatory development phase. Excludes future-trial mentions. | *"Phase I trial", "Phase 1/2"* | Other |
| No Trial-Specific Language Identified | No pattern above matched. Does not necessarily indicate absence of a trial component; for Clinical Trial Required projects, more likely reflects a limitation of abstract-based classification. | — | No Trial-Specific Language Identified |

### eTable 2. Manual overrides applied after 3 rounds of iterative review (n=34).

| Project Number | Activity | Reviewed in | Automated Classification (V3, Pre-Override) | Final Category (Override) | Rationale |
|---|---|---|---|---|---|
| 1K01MH132899-01A1 | K01 | Round 1 | No Trial-Specific Language Identified | Efficacy/Effectiveness | V3 automated output (No Trial-Specific Language Identified) unchanged from V2; set to manually reviewed assessment (Efficacy/Effectiveness). "We will compare the effects of education about herd immunity across these groups with the goal of identifying differential efficacy...” |
| 1K23DK140614-01A1 | K23 | Round 1 | Trial Confirmed, Stage Unclear | Other | V3 automated output (Trial Confirmed, Stage Unclear) unchanged from V2; set to manually reviewed assessment (Other). Mechanistic in nature based on hypothesis and aims |
| 1K23NS140632-01A1 | K23 | Round 1 | Trial Confirmed, Stage Unclear | Efficacy/Effectiveness | V3 automated output (Trial Confirmed, Stage Unclear) unchanged from V2; set to manually reviewed assessment (Efficacy/Effectiveness). Pilot was incorrectly assigned because of "I will first perform...” |
| 5K01AG075252-03 | K01 | Round 1 | Trial Confirmed, Stage Unclear | Efficacy/Effectiveness | V3 automated output (Trial Confirmed, Stage Unclear) unchanged from V2; set to manually reviewed assessment (Efficacy/Effectiveness). "...as well as determining the effects of...” |
| 5K01HL169495-02 | K01 | Round 1 | Trial Confirmed, Stage Unclear | Efficacy/Effectiveness | V3 automated output (Trial Confirmed, Stage Unclear) unchanged from V2; set to manually reviewed assessment (Efficacy/Effectiveness). "Aim 3 will evaluate whether Sleep Wizard is associated with improvements in...” |
| 5K01MH120321-05 | K01 | Round 1 | Trial Confirmed, Stage Unclear | Efficacy/Effectiveness | V3 automated output (Trial Confirmed, Stage Unclear) unchanged from V2; set to manually reviewed assessment (Efficacy/Effectiveness). "I will apply MOST and conduct a factorial optimization trial to isolate effects of...” |
| 5K08CA271949-04 | K08 | Round 1 | Pilot/Feasibility/Proof-of-Concept | Trial Confirmed, Stage Unclear | V3 automated output (Pilot/Feasibility/Proof-of-Concept) unchanged from V2; set to manually reviewed assessment (Trial Confirmed, Stage Unclear). "Develop a pilot" (not conduct a pilot trial). |
| 5K08CA273684-02 | K08 | Round 1 | Trial Confirmed, Stage Unclear | Pilot/Feasibility/Proof-of-Concept | V3 automated output (Trial Confirmed, Stage Unclear) unchanged from V2; set to manually reviewed assessment (Pilot/Feasibility/Proof-of-Concept). "The objective of this proposal is to evaluate feasibility of...” |
| 5K08HS029210-03 | K08 | Round 1 | Efficacy/Effectiveness | Pilot/Feasibility/Proof-of-Concept | V3 automated output (Efficacy/Effectiveness) unchanged from V2; set to manually reviewed assessment (Pilot/Feasibility/Proof-of-Concept). "establishing preliminary..." |
| 5K23DA053989-06 | K23 | Round 1 | Trial Confirmed, Stage Unclear | Efficacy/Effectiveness | V3 automated output (Trial Confirmed, Stage Unclear) unchanged from V2; set to manually reviewed assessment (Efficacy/Effectiveness). Aims are very clear about what the effectiveness and implementation outcomes. |
| 5K23HL150229-05 | K23 | Round 1 | Trial Confirmed, Stage Unclear | Pilot/Feasibility/Proof-of-Concept | V3 automated output (Trial Confirmed, Stage Unclear) unchanged from V2; set to manually reviewed assessment (Pilot/Feasibility/Proof-of-Concept). "The objectives of the proposed research are...evaluate feasibility and effectiveness of an early melatonin and sleep management intervention started during hospitalization...” |
| 5K23HL157765-04 | K23 | Round 1 | Efficacy/Effectiveness | Trial Confirmed, Stage Unclear | V3 automated output (Efficacy/Effectiveness) unchanged from V2; set to manually reviewed assessment (Trial Confirmed, Stage Unclear). "Dr. ... will perform a prospective study of implementation and effectiveness...” |
| 5K23HL159292-06 | K23 | Round 1 | Trial Confirmed, Stage Unclear | Efficacy/Effectiveness | V3 automated output (Trial Confirmed, Stage Unclear) unchanged from V2; set to manually reviewed assessment (Efficacy/Effectiveness). "MANATEE-T1D is a randomized double-blind placebo controlled trial...” |
| 5K23HL171940-02 | K23 | Round 1 | Trial Confirmed, Stage Unclear | Efficacy/Effectiveness | V3 automated output (Trial Confirmed, Stage Unclear) unchanged from V2; set to manually reviewed assessment (Efficacy/Effectiveness). Both aims are effectiveness and implementation. |

| Project Number | Activity | Reviewed in | Automated Classification (V3, Pre-Override) | Final Category (Override) | Rationale |
|---|---|---|---|---|---|
| 7K23DA062104-02 | K23 | Round 1 | Trial Confirmed, Stage Unclear | Efficacy/Effectiveness | V3 automated output (Trial Confirmed, Stage Unclear) unchanged from V2; set to manually reviewed assessment (Efficacy/Effectiveness). "Aim 2 will collect original data to examine changes ...during a JITAI RCT...” |
| 1K23HL175207-01A1 | K23 | Round 2 | Trial Confirmed, Stage Unclear | Pilot/Feasibility/Proof-of-Concept | V3 automated output (Trial Confirmed, Stage Unclear) unchanged from V2; set to manually reviewed assessment (Pilot/Feasibility/Proof-of-Concept). "...demonstrating the feasibility of a biomarker-driven clinical trial...” |
| 3K23HD098289-04S1 | K23 | Round 2 | Trial Confirmed, Stage Unclear | Pilot/Feasibility/Proof-of-Concept | V3 automated output (Trial Confirmed, Stage Unclear) unchanged from V2; set to manually reviewed assessment (Pilot/Feasibility/Proof-of-Concept). "...data will provide important information for development of a recruitment/retention toolkit to be supported through an anticipated R21 mechanism and a subsequent R01...” |
| 5K01HD106070-04 | K01 | Round 2 | No Trial-Specific Language Identified | Pilot/Feasibility/Proof-of-Concept | V3 automated output (No Trial-Specific Language Identified) unchanged from V2; set to manually reviewed assessment (Pilot/Feasibility/Proof-of-Concept). “The proposed program will adapt and pilot a ...” |
| 5K01HL163254-03 | K01 | Round 2 | Efficacy/Effectiveness | Pilot/Feasibility/Proof-of-Concept | V3 automated output (Efficacy/Effectiveness) unchanged from V2; set to manually reviewed assessment (Pilot/Feasibility/Proof-of-Concept). "The refined program will be tested in a fully powered effectiveness R01 trial submitted in Year 4..." |
| 5K01OH012795-02 | K01 | Round 2 | Trial Confirmed, Stage Unclear | No Trial-Specific Language Identified | V3 automated output (Trial Confirmed, Stage Unclear) unchanged from V2; set to manually reviewed assessment (No Trial-Specific Language Identified). Murine model, no text about randomizing, pilot, feasibility, etc. |
| 5K23AR079056-05 | K23 | Round 2 | No Trial-Specific Language Identified | Efficacy/Effectiveness | V3 automated output (No Trial-Specific Language Identified) unchanged from V2; set to manually reviewed assessment (Efficacy/Effectiveness). Significant detail in abstract describing the study of women, trial timelines, outcomes, and randomization groups/procedures. |
| 5K23DA054004-05 | K23 | Round 2 | Trial Confirmed, Stage Unclear | Efficacy/Effectiveness | V3 automated output (Trial Confirmed, Stage Unclear) unchanged from V2; set to manually reviewed assessment (Efficacy/Effectiveness). Very clear discussion on the design. |
| 5K23DK123416-05 | K23 | Round 2 | No Trial-Specific Language Identified | Trial Confirmed, Stage Unclear | V3 automated output (No Trial-Specific Language Identified) unchanged from V2; set to manually reviewed assessment (Trial Confirmed, Stage Unclear). "She will evaluate the intervention using a match cohort analysis..." |
| 5K23HL173684-02 | K23 | Round 2 | Pilot/Feasibility/Proof-of-Concept | Efficacy/Effectiveness | V3 automated output (Pilot/Feasibility/Proof-of-Concept) unchanged from V2; set to manually reviewed assessment (Efficacy/Effectiveness). Reference to prior feasibility/pilot before this award. “This career development award proposal includes a type 1 hybrid stepped-wedge cluster randomized trial among 4 sites and 900 women.” |
| 7K01MD018417-03 | K01 | Round 2 | No Trial-Specific Language Identified | Trial Confirmed, Stage Unclear | V3 automated output (No Trial-Specific Language Identified) unchanged from V2; set to manually reviewed assessment (Trial Confirmed, Stage Unclear). "I seek to develop and evaluate an intervention to improve...” |
| 1K01DA061444-01A1 | K01 | Round 3 | Other | Efficacy/Effectiveness | Round 3 manual review disagreement: "Phase 2" refers to a portion of the study (Phase 1 then Phase 2 of the study), not a regulatory Clinical Trial Phase II designation. “Phase 2 will use a double-blinded randomized crossover design where each participant will complete...” |
| 3K01DC019421-05S1 | K01 | Round 3 | No Trial-Specific Language Identified | Trial Confirmed, Stage Unclear | Round 3 manual review disagreement: Trial is conducted in humans, with functional MRI collected at multiple timepoints, confirming a human trial design. |

| Project Number | Activity | Reviewed in | Automated Classification (V3, Pre-Override) | Final Category (Override) | Rationale |
|---|---|---|---|---|---|
| 5K01AA028831-05 | K01 | Round 3 | Other | Trial Confirmed, Stage Unclear | Round 3 manual review disagreement: "Phase I" and "Phase II" language refers to parts/stages of the study itself, not regulatory Clinical Trial Phase I/II designation. |
| 5K01AR079043-04 | K01 | Round 3 | Efficacy/Effectiveness | Trial Confirmed, Stage Unclear | Round 3 manual review disagreement: No details on trial design/protocol/randomization; abstract only states they will evaluate effectiveness generically. “Phase II will involve a...two-group RCT...that compares...” |
| 5K08HS026530-06 | K08 | Round 3 | No Trial-Specific Language Identified | Pilot/Feasibility/Proof-of-Concept | Round 3 manual review disagreement: "Designing and evaluating provider-facing behavioral interventions to influence prescribing practices" indicates pilot/feasibility-stage. |
| 5K23HL151758-05 | K23 | Round 3 | Trial Confirmed, Stage Unclear | Efficacy/Effectiveness | Round 3 manual review disagreement: "...a randomized double-blinded sequential crossover study of...to determine...” |
| 5K23MH122777-04 | K23 | Round 3 | Pilot/Feasibility/Proof-of-Concept | No Trial-Specific Language Identified | Round 3 manual review disagreement: The "pilot trial" reference is to a different, independently funded project with data collection occurring in the past; the K award will evaluate that independently funded project. |
| 5K23MH135213-02 | K23 | Round 3 | No Trial-Specific Language Identified | Trial Confirmed, Stage Unclear | Round 3 manual review disagreement: "The current project will use a randomized parallel-group experimental therapeutic design to explore...". |
| 7K23AA028238-05 | K23 | Round 3 | No Trial-Specific Language Identified | Trial Confirmed, Stage Unclear | Round 3 manual review disagreement: Abstract states "we will recruit N=61 heavy drinking regular users..." confirming a human study with a defined recruitment target. |

For projects manually reviewed for Versions 1 and 2, if Version 3's automated classification was unchanged from Version 2's automated classification, the project's final category was set to the reviewer's manual assessment; otherwise Version 3's automated output was retained. For projects manually reviewed in Round 3 (evaluated directly against Version 3), any disagreement between the automated classification and manual assessment was overridden to the manual assessment directly. The rows below reflect the cumulative set of manual overrides across all 3 review rounds.

**eTable 3. Funding Institute/Center for K01/K08/K23 Awards Funded Under Clinical Trial Required and Clinical Trial Optional FOAs, FY2024-2025 (Full Breakdown)**

| Funding IC | Total (N=1336) | CT Required (N=1051) | CT Optional (N=285) |
|---|---|---|---|
| NHLBI | 236 (17.7) | 201 (19.1) | 35 (12.3) |
| NIMH | 169 (12.6) | 147 (14.0) | 22 (7.7) |
| NIDDK | 169 (12.6) | 135 (12.8) | 34 (11.9) |
| NIDA | 125 (9.4) | 111 (10.6) | 14 (4.9) |
| NCI | 112 (8.4) | 100 (9.5) | 12 (4.2) |
| NIA | 95 (7.1) | 78 (7.4) | 17 (6.0) |
| NICHD | 66 (4.9) | 64 (6.1) | 2 (0.7) |
| AHRQ | 64 (4.8) | 0 (0.0) | 64 (22.5) |
| NIMHD | 56 (4.2) | 47 (4.5) | 9 (3.2) |
| NIAAA | 41 (3.1) | 34 (3.2) | 7 (2.5) |
| NINDS | 34 (2.5) | 25 (2.4) | 9 (3.2) |
| NCCIH | 32 (2.4) | 32 (3.0) | 0 (0.0) |
| NIAMS | 30 (2.2) | 23 (2.2) | 7 (2.5) |
| OD | 27 (2.0) | 13 (1.2) | 14 (4.9) |
| FIC | 13 (1.0) | 12 (1.1) | 1 (0.4) |
| NIOSH | 12 (0.9) | 0 (0.0) | 12 (4.2) |
| NIDCD | 12 (0.9) | 10 (1.0) | 2 (0.7) |
| NIAID | 12 (0.9) | 0 (0.0) | 12 (4.2) |
| NINR | 11 (0.8) | 10 (1.0) | 1 (0.4) |
| NIGMS | 5 (0.4) | 3 (0.3) | 2 (0.7) |
| NIDCR | 5 (0.4) | 4 (0.4) | 1 (0.4) |
| NIEHS | 4 (0.3) | 2 (0.2) | 2 (0.7) |
| NEI | 4 (0.3) | 0 (0.0) | 4 (1.4) |
| NHGRI | 1 (0.1) | 0 (0.0) | 1 (0.4) |
| NIBIB | 1 (0.1) | 0 (0.0) | 1 (0.4) |

Values are n (column %) unless otherwise noted.

*Abbreviations:* AHRQ, Agency for Healthcare Research and Quality; CT, clinical trial; FIC, Fogarty International Center; NCCIH, National Center for Complementary and Integrative Health; NCI, National Cancer Institute; NEI, National Eye Institute; NHGRI, National Human Genome Research Institute; NHLBI, National Heart, Lung, and Blood Institute; NIA, National Institute on Aging; NIAAA, National Institute on Alcohol Abuse and Alcoholism; NIAID, National Institute of Allergy and Infectious Diseases; NIAMS, National Institute of Arthritis and Musculoskeletal and Skin Diseases; NIBIB, National Institute of Biomedical Imaging and Bioengineering; NICHD, Eunice Kennedy Shriver National Institute of Child Health and Human Development; NIDA, National Institute on Drug Abuse; NIDCD, National Institute on Deafness and Other Communication Disorders; NIDCR, National Institute of Dental and Craniofacial Research; NIDDK, National Institute of Diabetes and Digestive and Kidney Diseases; NIEHS, National Institute of Environmental Health Sciences; NIGMS, National Institute of General Medical Sciences; NIMH, National Institute of Mental Health; NIMHD, National Institute on Minority Health and Health Disparities; NINDS, National Institute of Neurological Disorders and Stroke; NINR, National Institute of Nursing Research; NIOSH, National Institute for Occupational Safety and Health; OD, Office of the Director.
